# Prevalence of carbapenem-resistant and extended-spectrum beta-lactamase-producing Enterobacteriaceae in a teaching hospital in Ghana

**DOI:** 10.1101/2022.08.24.22279171

**Authors:** James Sampah, Isaac Owusu-Frimpong, Frank T. Aboagye, Alex Owusu-Ofori

**Affiliations:** Department of Clinical Microbiology, School of Medicine and Dentistry, Kwame Nkrumah University of Science and Technology, Kumasi, Ghana; Biomedical and Public Health Research Unit, CSIR-Water Research Institute, Accra, Ghana; Clinical Microbiology Unit, Laboratory Services Directorate Komfo Anokye Teaching Hospital, Kumasi, Ghana; Laboratory Department, St. Patrick’s Hospital, Offinso, Ghana

**Keywords:** Antibiotic resistance, extended-spectrum beta-lactamases, carbapenem resistance, Enterobacteriaceae

## Abstract

**Background:** Carbapenem-resistant *Enterobacteriaceae* (CRE) and Extended-spectrum beta-lactamase (ESBL) production among Gram-negative Enterobacteriaceae is an increasing global challenge because of the high morbidity and mortality associated with their infections, especially in developing countries where there may not be many antibiotic treatment options. Despite these challenges, there are few studies in Ghana that have described the burden of CRE. This study aimed to determine the prevalence of carbapenem-resistant Enterobacteriaceae isolated from patients at the Cape Coast Teaching Hospital (CCTH) in the Central region of Ghana.

**Methodology/Principal findings:** Enterobacteriaceae isolates were collected from April to July 2019 at the bacteriology unit of CCTH using a consecutive sampling method. Isolates were identified by standard microbiological techniques and confirmed using API 20E. Kirby Bauer disc diffusion method was used to determine the antibiogram of isolates. Isolates were also subjected to ESBL testing using the single-disc combination method. Carbapenem resistant isolates were identified by the Kirby Bauer disc diffusion method and then subjected to genetic confirmation using polymerase chain reaction (PCR). Of the 230 isolates comprising *E. coli* (40.9%), *Citrobacter spp*. (32.6%), *K. pneumoniae* (9.1%), *P. mirabilis* (6.1%), *P. vulgaris* (5.2%), *Enterobacter spp* (3.5%)., *K. oxytoca* (2.2%), and *Serratia marcenses* (0.4%). Majority of the isolates were from urine 162(70.4%) and wound samples. The isolates showed high resistance to ampicillin 171 (74.3%) and cefuroxime 134(58.3%). The prevalence of MDR was 35.2% (81) with *E. coli* 40(42.6%) being the majority that exhibited MDR. Out of the 230 isolates, 113(49.1%) were ESBL producers with *E. coli* 54(57.5%) accounting for the majority while *Serratia marcenses* was the least. Out of the 13 (5.7%) CRE isolates which showed resistance towards carbapenem in the disc diffusion method, 11 expressed the blaNDM-1 gene whilst all isolates which showed resistant CRE expressed the blaOXA-48 gene.

**Conclusion:** The prevalence of carbapenem resistance and ESBL- producing Enterobacteriaceae pathogens among patients at the CCTH, Cape coast of Ghana is high, and effective infection prevention and control measures should be implemented at the hospital to prevent the rapid spread of these dangerous organisms.

## Introduction

The occurrence of antimicrobial resistance (AMR) is a significant challenge in the treatment of infections and these infectious diseases are ever-increasing especially with the re-emergence of pathogens [1]. AMR is considered to occur naturally and has been known to be escalated by the misuse and overuse of antimicrobial agents [2]. The increasing emergence of multidrug-resistant (MDR) Gram-negative bacilli is of great priority in clinical settings across the globe. Management of Gram-negative multidrug-resistant (MDR) infections has become significantly challenging over the past two decades in many developing countries, especially within the Sub-Saharan region [3]. These infections are usually associated with high morbidity rates, high mortalities, and extended hospital stays [4].

Enterobacteriaceae account for more than 30% of bacterial infections with high morbidity and mortality outcomes [5, 6]. Meningitis, urinary tract infections, gastroenteritis, septicemia, pneumonia, and wound infections are just a few of the infections that these organisms commonly cause [7, 8]. Enterobacteriaceae are well known for their public health threat across the globe due to their increasing antimicrobial resistance. Studies have established that members of this family of bacteria gain their antimicrobial resistance via the acquisition of drug-resistance genes through mobile genetic elements such as transposons and plasmids transferred within the same species or different species [9, 10]. The acquired resistance genes facilitate the production of β-lactamase enzymes, especially the extended-spectrum β-lactamase (ESBL), responsible for conferring resistance to the majority of β-lactam antibiotics [7, 10–13]. For instance, carbapenems, a class of β-lactam antibiotics, have been established to lose their potency against Enterobacteriaceae due to resistance [7, 14]. Other well-known antimicrobials such as fluoroquinolones, aminoglycosides, phenicols, sulfonamides, and tetracyclines have been rendered ineffective by this group of bacteria, thereby making treatment of infections of these bacteria difficult [15–17].

Carbapenems are broad-spectrum β-lactam antibiotics that are globally regarded as the ‘last-line’ antibiotics, thus, they are considered the last choice of drug for the treatment of critically ill patients and/or those infected with resistant Gram-negative bacteria [18]. They are essentially reserved for cases of suspected MDR bacterial infections. This class of β-lactam is very similar to penicillin and cephalosporin [19]. Carbapenems are bactericidal in their mode of activity against bacterial species that are Gram-negative. Unlike other β-lactam antibiotics, carbapenems invade the bacterial cell through the outer membrane proteins (*OprDs*), other than those used by the cephalosporin and penicillin (*OmpC* and *OmpF*), which results in the interruption of cell wall formation [18]. Once the cell wall formation is interrupted by the carbapenems, the peptidoglycan layer becomes very weak and the cell eventually bursts leading to the death of the bacterial cell [20, 21].

Generally, carbapenem-resistant Enterobacteriaceae (CRE) has been studied and reported in a few African countries such as Tanzania, South Africa, Nigeria, Kenya, Morocco, and Ghana [14, 22–36]. In Ghana, reported carbapenem resistance is not too different from other developing and under-developed countries in Africa. In a study by Codjoe and colleagues among Gram-negative isolates collected from four hospitals in Ghana, a 2.9% prevalence of carbapenem-resistant was reported [37]. The report further indicated that 23.4 % of the bacterial isolates were harbouring known carbapenem-resistant genes; *bla VIM-1, bla Oxa-48*, and *bla NDM-1*. Another study conducted by Quansah and colleagues reported carbapenem-resistant genes *OXA-48* (2.16%) and *NDM-1* (0.72%) in the study population [36].

While there are a few studies in other parts of Ghana that have looked at carbapenem resistance in Enterobacteriaceae, there are no published data on CRE from the Central Region of Ghana. As result, this study aimed to determine the prevalence of carbapenem-resistance, MDR, and ESBL-producing Enterobacteriaceae in the Cape Coast Teaching Hospital, in the Central Region of Ghana.

## Materials and methods

### Ethics statement

Cape Coast Teaching Hospital Ethical Review Committee (CCTHERC/EC/2019/044) of the Cape Coast Teaching Hospital and Committee on Human Research, Publication and Ethics (CHRPE/AP/201/19) of the School of Medical Sciences, Kwame Nkrumah University of Science and Technology jointly approved the study. Informed consent was obtained from all participants either by thumbprint or signature after an explanation of the procedure and the purpose of the study was provided to the patient. Informed consent was obtained from parents or guardians for participants below 18yrs.

### Study setting

The study was conducted between April and October 2019, at Cape Coast Teaching Hospital (CCTH) in Cape Coast in the Cape Coast Metropolitan, in the Central region of Ghana (Fig 1). The health facility is a tertiary government healthcare facility with a bed capacity of 420. In 2018, the facility recorded out-patient attendance of 158,164 while the total admission was 10,865 [38]. During the period of sample collection (April to July 2019), the diagnostic bacteriology unit received and processed 1,388 clinical specimens [38]. CCTH serves as a referral hospital for the Central Region and other parts of the Western Region. The metropolitan has a recorded population of 169,894, which comprises 82,810 (48.7%) males and 87,084 (51.3%) females [39].

**Fig 1.**
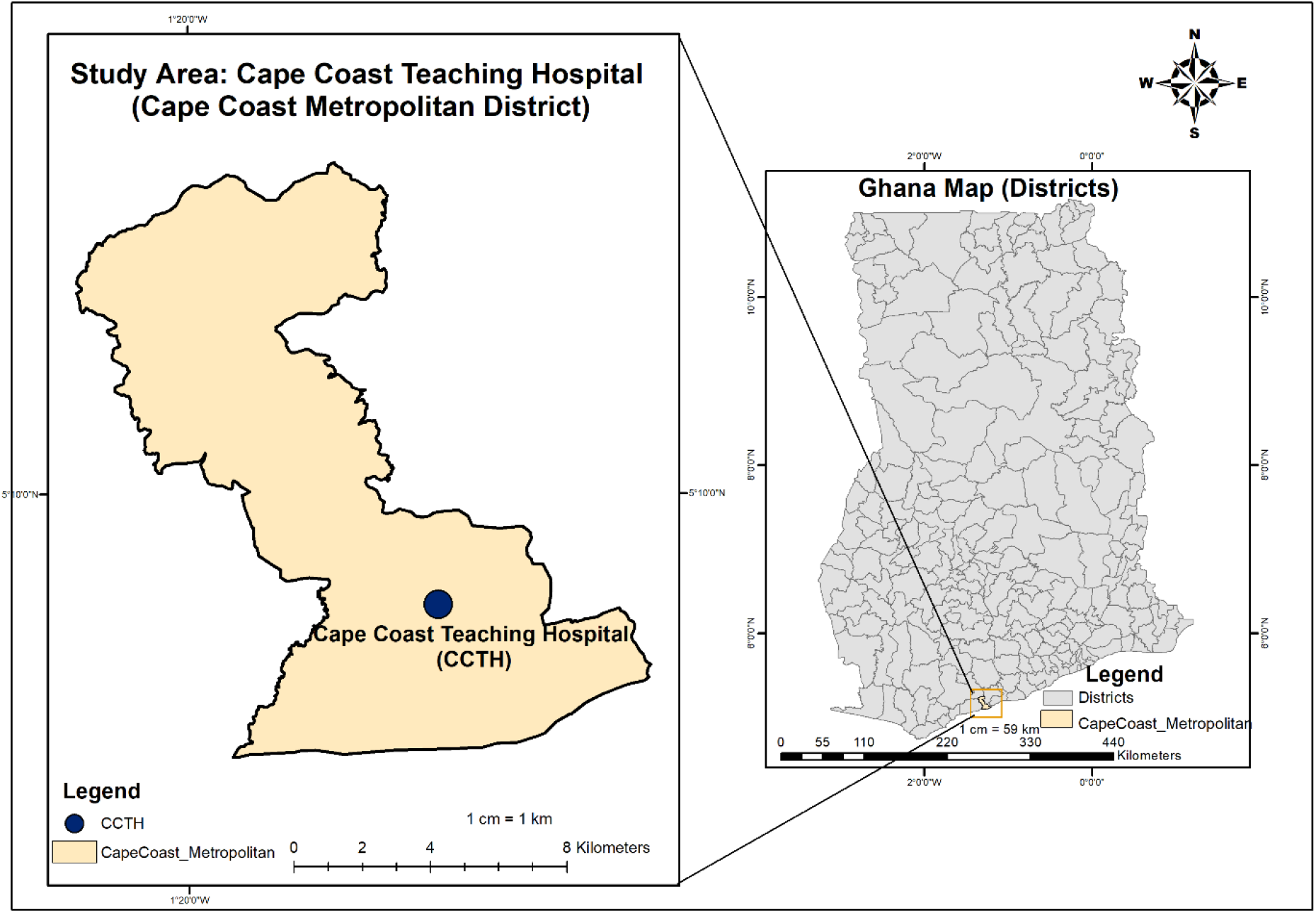
The map of Ghana showing the location of Cape Coast Teaching Hospital in the Cape Coast Metropolitan District. *Map developed with ESRI ArcMap 10*.*8 using data from Ghana Open Data Initiative, and OpenStreetMap and OpenStreetMap Foundation*.

### Bacterial collection

Two hundred and thirty (230) non-repetitive *Enterobacteriaceae* were isolated and identified from the clinical specimens submitted using standard bacteriological techniques and a panel of biochemical tests. Isolates were from urine, blood, sputum, high vaginal swab (HVS), endocervical swab, wound swab, stools, and semen presented to the bacteriology unit at CCTH. Using a pretested questionnaire, sociodemographic data including age and sex were recorded. Isolates identification was confirmed using the API 20E strip (BioMerieux, France). Isolates were maintained in 20% brain heart infusion glycerol broth at -20^0^C for further testing.

### Antibiotic susceptibility testing

All isolates from glycerol broth were subcultured on MacConkey agar media after partially thawing the broth. Antibiotic susceptibility testing (AST) was performed on Mueller Hinton agar using the Kirby Bauer disc diffusion method and interpreted according to the Clinical and Laboratory Standards Institute (CLSI) guideline [40]. The AST was performed using the following antibiotic discs; Ampicillin (10µg), Ciprofloxacin (5µg), Levofloxacin (5µg), Ofloxacin (5µg), Gentamicin (10µg), Amikacin (30µg), Ceftriaxone (30µg), Ceftazidime (30µg), Cefotaxime (30µg) and Cefuroxime (30µg) which were from Biomark Laboratories, India. Antibiotic discs Meropenem (10µg), Imipenem (10µg), and Ertapenem (10µg) from Oxoid, UK were included. *E. coli* (NCTC 19418) was used as a quality control strain. The zones of inhibitions were recorded and interpreted according to the Clinical and Laboratories Standards Institute guidelines [40]. Multidrug resistance was defined as resistance to at least three classes of antibiotics [21].

### Phenotypic screening and confirmation for ESBL

ESBL detection was carried out on Mueller Hinton agar seeded with the test organism. All 230 isolates were screened for ESBL-producing enzymes using Ceftazidime (30µg) and Cefotaxime (30µg) discs according to the method described by the CLSI guideline [40]. An isolate resistant to any of the screening antibiotic discs was suspected of ESBL and reported as positive for ESBL screening. ESBL confirmation was done using a single disc of Oxoid Cefpodoxime (10µg) alone and Cefpodoxime/ clavulanic acid (10/1µg) using the Kirby Bauer disc diffusion method [40]. The confirmatory discs were placed on the seeded Mueller Hinton agar ensuring that the discs were at least 20mm apart and incubated at 37^0^C overnight. An enhanced zone of inhibition (≥ 5 mm) around the Cefpodoxime/ clavulanic acid (10/1µg) relative to the single disc of Cefpodoxime (10µg) was considered positive for the production of ESBL. *Escherichia Coli* ATCC 25299 and *K. pneumoniae* ATCC 700603 were used as quality controls.

### Molecular detection of carbapenem-resistant genes

After the antibiotic susceptibility testing using antibiotic discs, isolates that were resistant and intermediate susceptible to ertapenem, meropenem or imipenem were selected for genetic confirmation using polymerase chain reaction (PCR).

### Bacterial DNA extraction

The DNA of bacteria isolates was extracted using Quick DNA kits (Zymo Research, USA) according to the manufacturer’s procedure. Using a sterile loop, a loopful of the isolates were picked from the Mueller Hinton agar plate and emulsified in sterile 1.5 mL microcentrifuge tubes containing 400 µL of Genomic Lysis Buffer and 5 µL Proteinase K. These tubes were vortexed at 2500 rpm for 30sec and incubated at 56°C overnight. After the overnight incubation, mixtures were vortexed again and transferred to Zymo-Spin™ IIC Columns in collection tubes and centrifuged at 10,000 x g for one minute. The flow-throughs were discarded, after which 200 µL of DNA Pre-Wash Buffer was added to each spin column. Centrifugation was performed again at 10,000 x g for one minute. Afterward, the final washing was done by adding 500 μL of g-DNA Wash Buffer to the spin column and centrifuging again at 10,000 x g for one minute. Finally, the spin columns were transferred into sterile 1.5 mL microcentrifuge tubes, after which 100 μL of Elution Buffer was added to elute the DNA. DNA samples were stored at -20ºC before proceeding to downstream analysis. DNA quantification was performed to determine the individual concentrations of the isolate with the Qubit 3.0 fluorometer (Life Technologies Holdings Pte Ltd, Malaysia) and the values recorded.

### Detection of carbapenem resistance-encoding genes using PCR

Carbapenem-resistant isolates were examined for the presence of *blaKPC-1, blaIMP-1, blaVIM-1, blaNDM-1*, and *blaOXA-48* genes by PCR methods using gene-specific primers (Table 1) described by Poirel and colleagues [27]. The PCR assay was carried out with a 20 µL reaction mixture containing 2 µL genomic-DNA, 1x Standard reaction buffer, 0.3mM each of dATP, dCTP, dTTP, and dGTP, 200 nM each of Forward and Reverse primers, and 1.25 U One*Taq* DNA Polymerase (New England Biolabs Inc., USA). PCR reactions were performed using the Bio-Rad PTC-200 Thermal Cycler (Bio-Rad Laboratories, USA) with the following cycling conditions; an initial denaturation at 94 ^0^C for 3 mins, followed by 40 cycles at 94 ^0^C for 30sec, 56 ^0^C for 30sec, and 68 ^0^C for 30sec. Finally, an elongation step was performed at 68ºC for 5 minutes. Afterward, the PCR products were resolved by agarose gel electrophoresis and visualized under UV light using UVP Bio-Doc-It Imaging system – trans-illuminator (AnalytikJena, Germany).

**Table 1.**
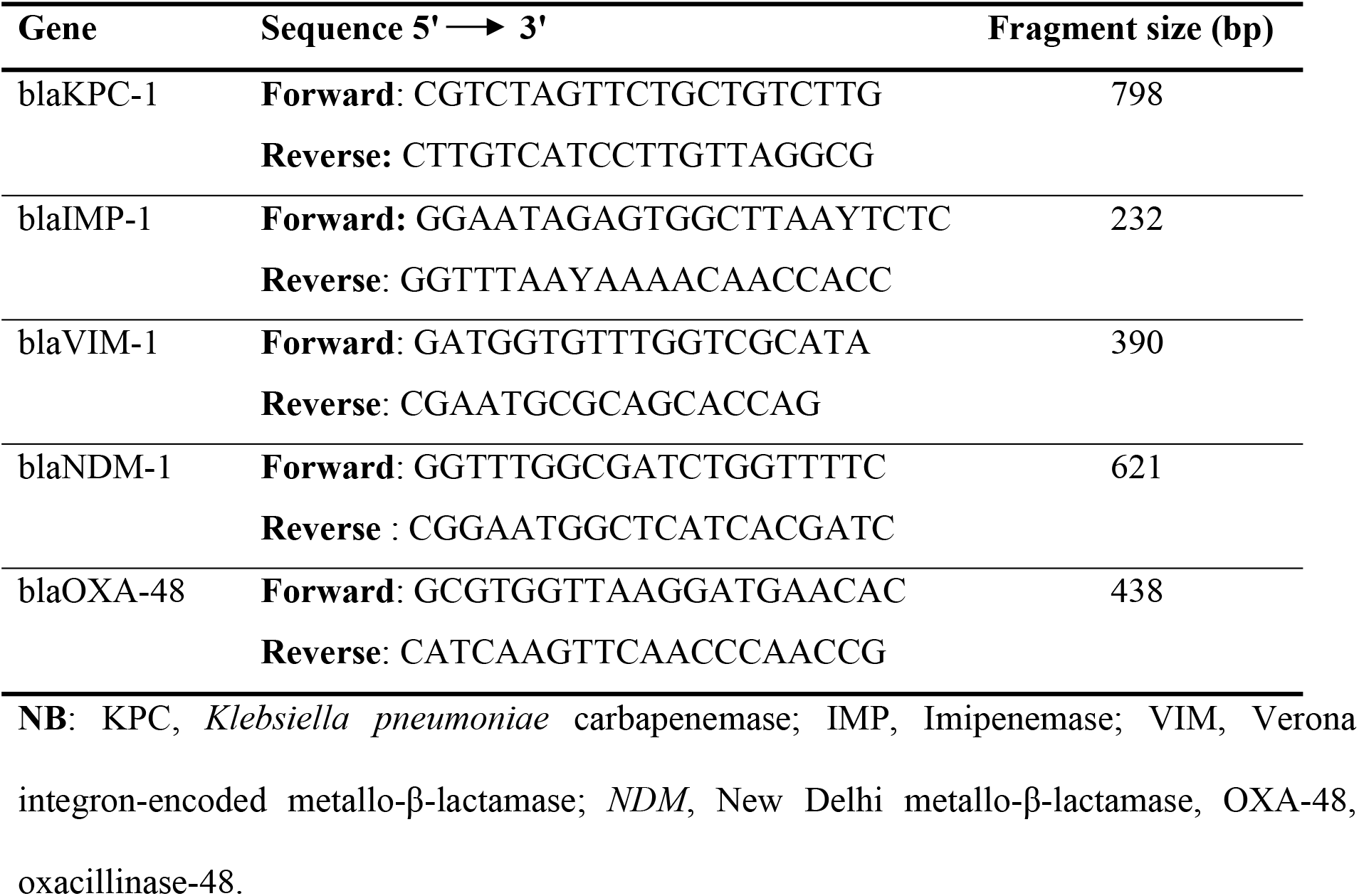
Carbapenem-resistant genes primer sets.

### Statistical analysis

Data were entered using Microsoft Excel 2019 and analyzed using GraphPad Prism version 8.0 (Graphpad Inc., La Jolla, CA, USA). A simple frequency was used to describe the study population with the socio-demographic and other relevant variables.

## Results

### Distribution of clinical specimen

Out of the 1388 samples processed, 230 isolates were identified as Enterobacteriaceae; 162 (70.4%) isolates were from urine, 23 (10.0%) from wound swab, 22 (9.6%) from HVS, 15 (6.5%) from sputum, 4 (1.7%) from blood, 2 (0.8%) from endocervical swab, 1 (0.4%) from other types of specimens (Fig 2).

**Fig 2.**
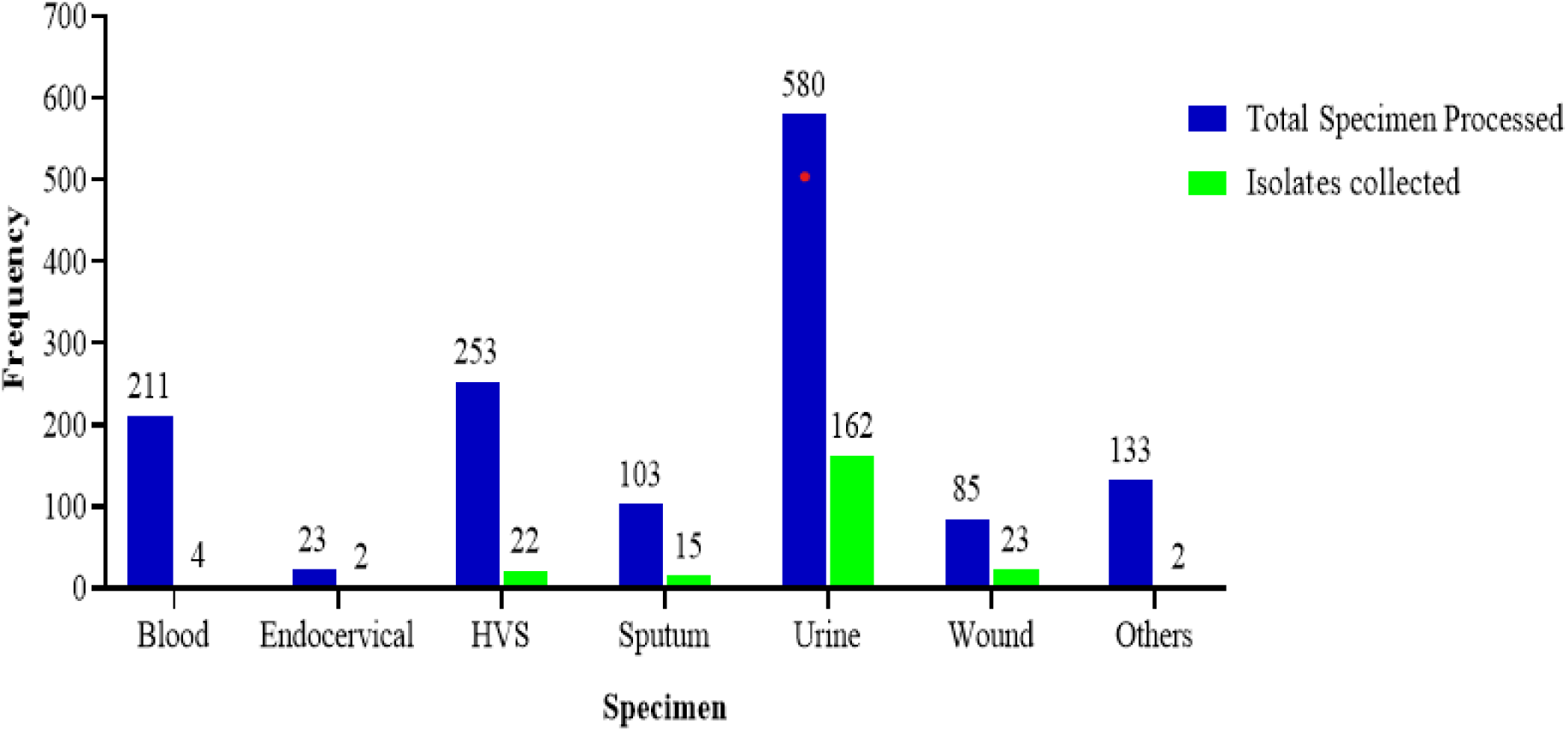
Distribution of samples received, and isolates collected Sociodemographic characteristics of the study population.

The study enrolled 230 participants from whose clinical specimens *Enterobacteriaceae* were isolated. Out of the total participants, 196 (85.2%) were out-patients while 34 (14.8%) were on admission at various wards. Most participants were >70years 43 (18.7%) while the age group with the least number of participants was children <10yrs 8 (3.5%). One hundred and sixty-six (166) females representing 72.2% participated in the study while 64 (27.8%) were males (Table 2).

**Table 2.** Gender and age distribution among participants.

|  | Age group [Number of participants (N)] |  |  |  |  |  |  |  | Total (%) |
| --- | --- | --- | --- | --- | --- | --- | --- | --- | --- |
|  | <10 | 11-20 | 21-30 | 31-40 | 41-50 | 51-60 | 61-70 | >70 |  |
| **Male** | 2 | 2 | 6 | 6 | 2 | 10 | 15 | 21 | 64 (27.8) |
| **Female** | 6 | 12 | 35 | 28 | 20 | 22 | 21 | 22 | 166 (72.2) |
| **Outpatient** | 3 | 12 | 32 | 33 | 20 | 26 | 33 | 37 | 196 (85.2) |
| **In-patients** | 5 | 2 | 9 | 1 | 2 | 6 | 3 | 6 | 34 (14.8) |

### Distribution of isolates/organism

Of the 230 isolates collected for this study, *Escherichia coli* was the most frequent pathogen 94 (40.9%), followed by *Citrobacter* spp. 75 (32.6%), *Klebsiella pneumoniae* 21 (9.1%), *Proteus vulgaris* 12 (5.2%), *Proteus mirabilis* 14 (6.1%), *Enterobacter* spp. 8 (3.5%) *Klebsiella oxytoca* 5 (2.2%) and *Serratia marcenses* 1 (0.4%) (Table 3).

**Table 3:** Distribution of isolates.

| Isolate | Point of Care |  |  |
| --- | --- | --- | --- |
|  | Outpatient [N] | In-patient [N] | Total [N] (%) |
| *E. coli* | 80 | 14 | 94 (40.9) |
| *Citrobacter* spp. | 66 | 9 | 75 (32.6) |
| *K. pneumoniae* | 19 | 2 | 21 (9.1) |
| *P. vulgaris* | 9 | 3 | 12 (5.2) |
| *P. mirabilis* | 8 | 6 | 14 (6.1) |
| *Enterobacter* spp. | 8 | 0 | 8 (3.5) |
| *K. oxytoca* | 5 | 0 | 5 (2.2) |
| *Serratia marcescens* | 1 | 0 | 1 (0.4) |
| **Frequency** | **196** | **34** | **230 (100)** |

### Antibiotics susceptibility pattern of Enterobacteriaceae

Out of the total of 13 antibiotics tested, susceptibility was highest to amikacin (AMK) (97.8%). This was followed by meropenem (MEM), imipenem (IMI), and ertapenem (ERT) with frequencies of 224 (97.4%), 224 (97.4%), and 204 (88.7%), respectively. The antibiotics to which isolates were most resistant were ampicillin (AMP) (74.4%), cefuroxime (CXM) (58.3%), and cefotaxime (CTX) (50.9%). Concerning the use of other antibiotics, isolates were resistant to Ciprofloxacin (33.0%) and gentamicin (19.2%) (Table 4). Penicillin (74.3%) was the class of antibiotic to which isolates showed the most resistance whilst carbapenem (3.6%) was the class of antibiotic to which isolates showed the least resistance (Fig 3).

**Table 4:** Antibiotic susceptibility profile of *Enterobacteriaceae*.

| Antibiotic | Susceptible *n* (%) | Intermediate *n* (%) | Resistant *n* (%) |
| --- | --- | --- | --- |
| Ampicillin | 46 (20) | 13 (5.7) | 171 (74.3) |
| Gentamicin | 156 (67.8) | 30 (13.0) | 44 (19.1) |
| Amikacin | 225 (97.8) | 3 (1.3) | 2 (0.9) |
| Ciprofloxacin | 141 (61.3) | 13 (5.7) | 76 (33.0) |
| Levofloxacin | 158 (68.7) | 12 (5.2) | 60 (26.1) |
| Ofloxacin | 145 (63) | 13 (5.7) | 72 (31.3) |
| Cefuroxime | 80 (34.8) | 16 (6.9) | 134 (58.3) |
| Cefotaxime | 104 (45.2) | 9 (3.9) | 115 (50.9) |
| Ceftazidime | 106 (46.1) | 9 (3.9) | 115 (50.0) |
| Ceftriaxone | 108 (47.0) | 9 (3.9) | 113 (49.1) |
| Ertapenem | 204 (88.6) | 13 (5.7) | 13 (5.7) |
| Meropenem | 224 (97.4) | 0 (0.0) | 6 (2.6) |
| Imipenem | 224 (97.4) | 0 (0.0) | 6 (2.6) |

**Fig 3.**
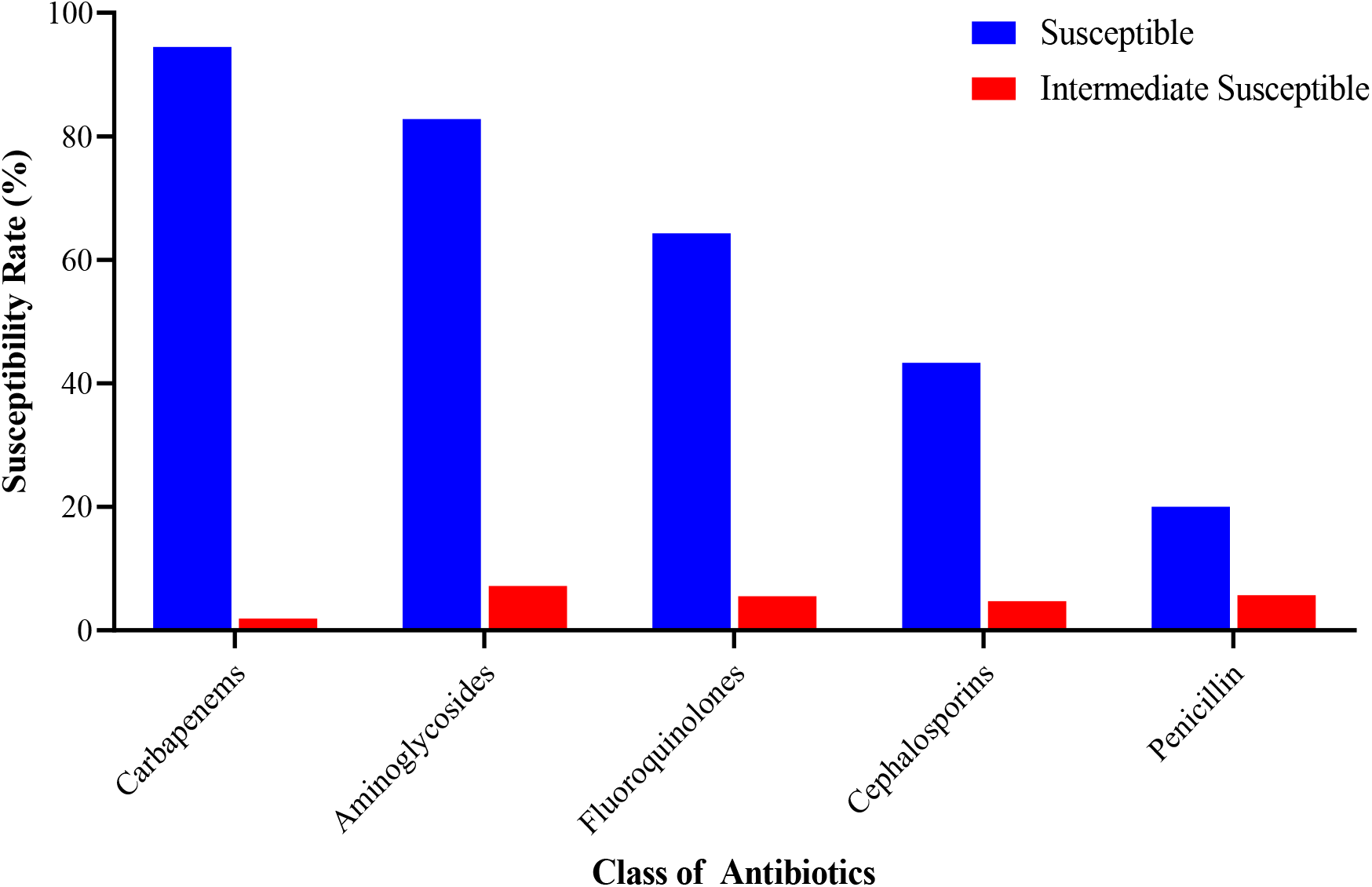
Susceptibility of isolates to the different classes of antibiotics.

### MDR and Extended-Spectrum Beta-Lactamase (ESBL) producing organisms

Out of the 230 isolates collected, eighty-one (81) isolates exhibited multidrug resistance (35.2%) toward antibiotics used. Among isolates that exhibited MDR, *E. coli* (42.6%) was in the majority while *Serratia marcenses* was the least. Out of 230 isolates screened for ESBL production, 113 (49.1%) were ESBL producers while 117 (50.9%) were non-ESBL producers. Among the ESBL producers, *E. coli* (23.5%) was in the majority while *Serratia marcenses* (0.4%) was the least. ESBL producers were found among all the species of isolates collected. (Table 5).

**Table 5:** MDR and ESBL among isolates.

| Bacterial isolates | Number of isolates (n) | MDR<br>n (%) | ESBL<br>n (%) | Non-ESBL<br>n (%) |
| --- | --- | --- | --- | --- |
| *E.coli* | 94 | 40 (42.6) | 54 (57.5) | 40 (42.6) |
| *Citrobacter* spp. | 75 | 25 (33.3) | 31 (41.3) | 44 (58.7) |
| *Klebsiella pneumoniae* | 21 | 5 (23.8) | 10 (47.6) | 11 (52.4) |
| *Proteus mirabilis* | 14 | 4 (28.6) | 7 (50.0) | 7 (50.0) |
| *Proteus vulgaris* | 12 | 3 (25.0) | 4 (33.3) | 8 (66.7) |
| *Enterobacter* spp. | 8 | 2 (25.0) | 3 (37.5) | 5 (62.5) |
| *Klebsiella oxytoca* | 5 | 1 (20.0) | 3 (60) | 2 (40.0) |
| *Serratia marcescens* | 1 | 1 (100) | 1 (100) | 0 (0.0) |
| **Frequency** | **230** | **81 (35.2)** | **113 (49.1)** | **117 (50.9)** |

### Molecular detection of Carbapenem-resistant genes

Thirteen (13) isolates that showed resistance and 13 isolates that showed intermediate resistance to carbapenem from the Kirby Bauer disc diffusion test were selected for PCR (Fig 4, A). None of the isolates selected for PCR expressed blaKPC-1, IMP-1, and VIM-1. BlaOXA-48 and blaNDM-1 were detected among some of the Carbapenem-Resistant Enterobacteriaceae (CRE) isolates (Fig 4, A). Out of the 13 resistant isolates, 11 expressed the blaNDM-1 gene whilst all resistant CRE expressed the blaOXA-48 gene (Fig 4, D). Also, 9 out of the 13 resistant isolates expressed both blaOXA-48 and blaNDM-1 resistant genes. *P. mirabilis* (26.7%) was the Enterobacteriaceae which expressed more of the OXA-48 gene whilst *E. coli* (3.2%) was the least among the isolates which expressed the OXA-48 gene (Fig 4, B). Also, *P. mirabilis* (20.0%) expressed more of the NDM-1 gene whilst *E. coli* (1.1%) was the isolate with the least expression of the NDM-1 gene (Fig 4, B). The 13 resistant isolates with carbapenem-resistant genes detected were found among both outpatients (53.8%) and in-patients (46.2%). The blaOXA-48 gene was also found among all 13 intermediate susceptible isolates (Fig 4, C).

**Fig 4.**
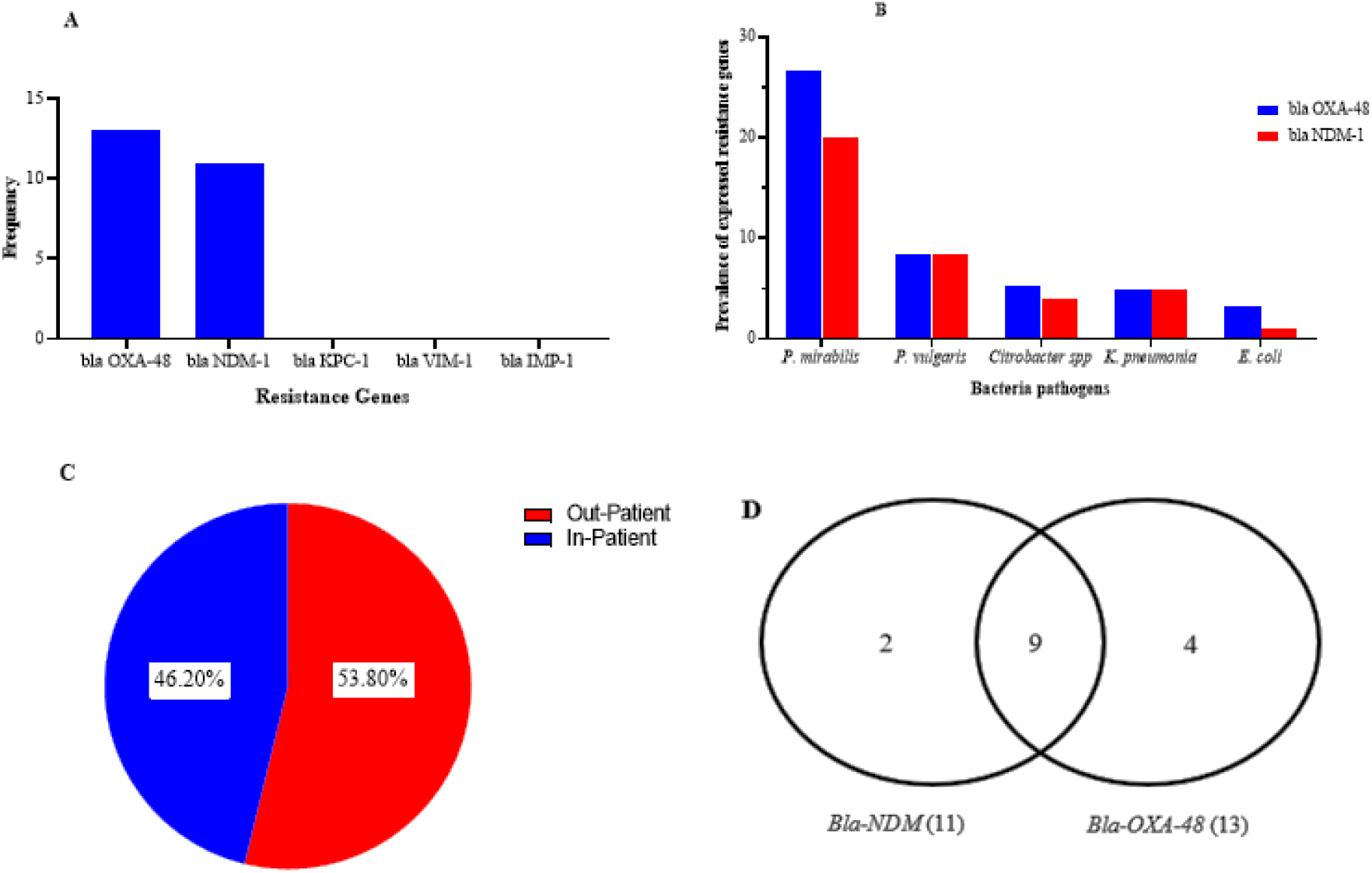
The outcome of Carbapenem-resistant genes from the molecular detection showing the presence of Carbapenem-resistant gene (A), distribution of bla OXA-48 and bla NDM-1 genes among CRE isolates (B), location of patients with CRE (C) and the distribution of *bla OXA-48* and *NDM-1* genes (D).

## Discussion

The knowledge of the distribution and surveillance of bacterial infections and their antibiotic profiles are very critical for the creation of awareness, implementation of infection control measures, and proper management of such infections. This is essential in developing countries, especially in sub-Saharan Africa where studies have shown that many health facilities have poor infection prevention and control (IPC) adherence. This challenge translates into increased bacterial infections caused by multi-drug resistant bacteria and subsequently resulted in increased morbidity and mortality [41, 42]. Resistance to *β*-lactam antimicrobials among Enterobacteriaceae has been caused largely by the acquisition of resistant genes that encodes for *β*-lactamase enzymes [43, 44].

From this study, the frequent isolates from clinical specimens were *Escherichia coli* (40.9%), *Citrobacter* spp (32.6%), and *Klebsiella pneumoniae* (9.1%). This finding agrees with the report by Blomberg and colleagues from a study conducted in Tanzania, which indicated that *Escherichia coli, Klebsiella oxytoca*, and *Klebsiella pneumoniae* are common bacteria isolated from clinical samples of patients [45]. The findings of this study are consistent with a finding in Ghana which reported *Klebsiella pneumoniae, Enterobacter spp*., and *Escherichia coli* as the most isolated pathogenic *Enterobacteriaceae* from clinical specimens [46]. Again, the results of the isolated organisms conform to the findings from a study conducted by Feglo and colleagues in Ghana which reported that the most isolated *Enterobacteriaceae* from clinical specimens were predominantly *E. coli, Klebsiella pneumoniae*, and *Klebsiella oxytoca* [47]. These commonly isolated bacteria have developed resistance to commonly used antibiotics thereby making it difficult to effectively treat infections caused by such bacteria [45].

Findings from this study have shown that *Citrobacter* spp (32.6%) is one of the common emerging uropathogens of urinary tract infections. Out of the 75 isolates of *Citrobacter* spp collected, 58(35.8%) of the total number of *Citrobacter* spp. were isolated from urine specimens. This agrees with the study conducted by Sami and colleagues which reported a prevalence of 3.5% *Citrobacter* spp [48], although this prevalence is lower than that of this study. The increase in the proportion of the *Citrobacter* spp. isolated could be attributed to the difference in geographical location.

The Enterobacteriaceae were predominantly isolated from females and mostly from urine specimens. The finding from this study is contrary to the study conducted by Nordmann and colleagues which indicated that there is no preference for sex by Enterobacteriaceae [49]. Their finding indicated that both genders are equally infected when the opportunity is created since these organisms are mainly opportunistic. Though the results from this study indicate that both sexes are affected, more females (166, 72.2%) were affected. The rate of antimicrobial drug resistance is increasing rapidly, and the majority of commonly used drugs are significantly becoming ineffective for antimicrobial therapy [49, 50]. Antimicrobial resistance (AMR) is a threat to the effective prevention and treatment of infectious diseases caused by infectious pathogens [1]. Across the globe, antibiotics are the mainstay in the effective control and management of bacterial infections in the clinical setting [21].

The susceptibility profile of Enterobacteriaceae isolates in this study showed a low multidrug resistance (32.5%) as compared to 89.5% in a similar study [4]. This study has shown a relatively low resistance toward ampicillin (74.4%), cefuroxime (58.3%), and cefotaxime (54.8%) as compared to the report by Agyepong and colleagues which indicated a high resistance of Gram-negative bacteria towards, ampicillin (94.4%), cefuroxime (79.0%) and cefotaxime (71.3%) [4]. Similarly, the results from this study were contrary to the report in Ghana by Feglo and colleagues which reported isolates resistant to ampicillin (91.7%) and cefuroxime (70.6%) [47]. Resistance of isolates towards cefotaxime (54.8%) was high in this study as compared to 48.1% as reported by Feglo and colleagues [47]. Results from the present study also support the findings by Labi and colleagues which indicated high antibiotic resistance among members of the *Enterobacteriaceae* at the Korle Bu Teaching Hospital to the combination of ampicillin/ gentamicin and ampicillin/ cefotaxime [51]. The resistance among *Enterobacteriaceae* to ampicillin and other commonly used antibiotics is consistent with other studies in sub-Saharan African countries such as Nigeria [52], Rwanda [53], Ethiopia [54], Zimbabwe [55], and Tanzania [56]. The trend of antibiotic resistance rates in the sub-region could be due to high antibiotic selection pressure which comes as a result of the unregulated availability of antibiotics over the counter and cheap substandard antibiotics influx in the sub-Saharan region [14, 57–59].

ESBL production by Gram-negative bacteria poses a great challenge in the management of Gram-negative bacterial infections. ESBL production may be associated with MDR. From the study, the prevalence of ESBL producers among *Enterobacteriaceae* was 49.1% (113/230) with the most ESBL-producing organism being *E. coli* (57.5%). The prevalence (49.1%) of the ESBL was lower than that reported by Feglo and colleagues in a study conducted in Ghana which reported a prevalence of 57.8% [47]. Furthermore, findings from this study showed that ESBL prevalence was higher than the ESBL prevalence (37.96 %) reported by Oduro-Mensah and colleagues in Ghana [60]. The difference in the prevalence could be attributed to the difference in geographical location, sample size, and the period of sampling. Also, the result on the ESBL producers indicated that the ESBL harbouring isolates were predominantly from outpatients (76.1%). This is not expected since inpatients are most likely to have nosocomial infections and have been exposed to more antibiotics so are likely to harbour ESBL producers. Inpatients are more at risk than outpatients. The ESBL distribution among outpatients and in-patients is contrary to the report by Khanfar and colleagues from Saudi Arabia which reported ESBL producers to be higher in inpatients (15.4%) than out-patients (4.5%) [61]. In like manner, a study conducted by Ouedraogo and colleagues in Burkina Faso in the sub-Saharan region also reported ESBL prevalence to be higher in hospitalized patients [62]. The high prevalence of ESBL producers among outpatients could be due to the possible self-medication due to the unregulated availability of antibiotics over the counter which may have escalated the transmission of the plasmid-encoded genes within the community. The community may serve as a reservoir for ESBL which may eventually result in a possible outbreak of ESBL-carrying strains of *Enterobacteriaceae* if not contained.

The outcome of the detection of the carbapenem-resistant gene via PCR showed the presence of *bla NDM-1* and *bla OXA-48* among different bacteria. In addition, the urine specimens exhibited more carbapenem-resistant genes than the other specimens. Also in this study, the most implicated bacteria were *Proteus* spp, *Citrobacter* spp, and *E. coli*. This result is in agreement with the report by Codjoe and colleagues [37]and Quansah and colleagues [36] which indicated the presence of both *bla NDM-1* and *bla OXA-48* in Ghana. Furthermore, the CRE-resistant isolates which harboured carbapenem-resistant genes were also ESBL-producing bacteria using the phenotypic method of detection. This agrees with the study conducted by Bornet and colleagues [63] which affirms that most CREs are ESBL producers. Reports from this study agree with surveillance studies in the western world and developed countries, which indicated that there had been reports of several rates of carbapenemase producers globally. In Europe, Germany had a prevalence of CRE of less than 1 [64]. When compared with prevalence in these developed countries, the prevalence from this study was high. This could be due to the well-regulated use of drugs (antibiotics) in developed countries compared to developing countries. For instance in Asia, China had a reported prevalence of 33.3% which was mainly due to *NDM-1-*producing CRE [65]. This prevalence in China is high when compared to the prevalence of CRE in this study. The possible importation of resistance genes has been found in a few countries such as South Africa due to tourism and migration [66].

## Conclusion

The study has demonstrated the high prevalence of carbapenem-resistant and high ESBL production among *Enterobacteriaceae*. ESBL production was found among 49.1% of isolates and the prevalence of MDR and CRE were 35.2% and 5.7% respectively. *E. coli, Citrobacter* spp., *K. pneumoniae*, and *Klebsiella oxytoca* were the most common causative organisms isolated. The study found high rates of MDR among *E. coli* and *Citrobacter* spp. The *Enterobacteriaceae* were most sensitive towards amikacin, imipenem, and meropenem and most resistant towards ampicillin, cefuroxime, and cefotaxime. These results should guide the empirical treatment of bacterial infections caused by *Enterobacteriaceae* in Cape Coast Teaching Hospital of Ghana. Other recommendations may include PCR to determine the types of ESBL in CCTH and adopting strict infection control measures to prevent the rapid spread of resistance.

## Data Availability

The data underlying the results presented in the study will be made available

## Acknowledgment

We would like to thank the Cape Coast Teaching Hospital staff, especially the technical staff at the bacteriology unit of the laboratory department.

## Availability of data and materials

Data and materials have been provided in the main manuscript. Additional information can be provided by the corresponding author upon request.

## Authors’ contributions

The study was conceptualized and jointly designed by JS and AOO. AOO supervised the work. JS and IOF collected the data and undertook laboratory analysis. Data analysis was done with assistance from FTA. JS, IOF, and FTA prepared the manuscript with the assistance of AOO. All authors read and approved the final manuscript.

## References

1. World Health Organization. Antimicrobial resistance:: Global report on surveillance; 2014.

2. World Health Organization 2018 Global Antimicrobial Resistance Surveillance System (GLASS) Report Early Implementation 2017–2018 2018.

3. Singh N, Manchanda V. Control of multidrug-resistant Gram-negative bacteria in low-and middle-income countries-high impact interventions without much resources. Clin Microbiol Infect. 2017;23:216–8. doi:10.1016/j.cmi.2017.02.034.

4. Agyepong N, Govinden U, Owusu-Ofori A, Essack SY. Multidrug-resistant gram-negative bacterial infections in a teaching hospital in Ghana. Antimicrob Resist Infect Control. 2018;7:37. doi:10.1186/s13756-018-0324-2.

5. Méndez-Vilas A. Microbial pathogens and strategies for combating them: Science, technology and education 2013;4. Badajoz: Formatex Research Center.

6. Rossolini GM, Mantengoli E, Docquier J-D, Musmanno RA, Coratza G. Epidemiology of infections caused by multiresistant gram-negatives: ESBLs, MBLs, panresistant strains. J Nat Sci. 2007;30:332–9.

7. Nordmann P, Naas T, Poirel L. Global spread of Carbapenemase-producing Enterobacteriaceae. Emerging Infectious Diseases. 2011;17:1791–8. doi:10.3201/eid1710.110655.

8. Paterson DL. Resistance in gram-negative bacteria: Enterobacteriaceae. American Journal of Infection Control. 2006;34:S20-8; discussion S64-73. doi:10.1016/j.ajic.2006.05.238.

9. Bitew A, Tsige E. High Prevalence of Multidrug-Resistant and Extended-Spectrum β-Lactamase-Producing Enterobacteriaceae: A Cross-Sectional Study at Arsho Advanced Medical Laboratory, Addis Ababa, Ethiopia. Journal of Tropical Medicine. 2020;2020:6167234. doi:10.1155/2020/6167234.

10. Shilpakar A, Ansari M, Rai KR, Rai G, Rai SK. Prevalence of multidrug-resistant and extended-spectrum beta-lactamase producing Gram-negative isolates from clinical samples in a tertiary care hospital of Nepal. Trop Med Health. 2021;49:23. doi:10.1186/s41182-021-00313-3.

11. Bradford PA. Extended-spectrum beta-lactamases in the 21st century: characterization, epidemiology, and detection of this important resistance threat. Clin Microbiol Rev. 2001;14:933-51, table of contents. doi:10.1128/CMR.14.4.933-951.2001.

12. Ben-Ami R, Rodríguez-Baño J, Arslan H, Pitout JDD, Quentin C, Calbo ES, et al. A multinational survey of risk factors for infection with extended-spectrum beta-lactamase-producing enterobacteriaceae in nonhospitalized patients. Clin Infect Dis. 2009;49:682–90. doi:10.1086/604713.

13. Perez F, van Duin D. Carbapenem-resistant Enterobacteriaceae: a menace to our most vulnerable patients. Cleveland Clinic Journal of Medicine. 2013;80:225–33. doi:10.3949/ccjm.80a.12182.

14. Mushi MF, Mshana SE, Imirzalioglu C, Bwanga F. Carbapenemase genes among multidrug resistant gram negative clinical isolates from a tertiary hospital in Mwanza, Tanzania. BioMed Research International. 2014;2014:303104. doi:10.1155/2014/303104.

15. Tada T, Miyoshi-Akiyama T, Dahal RK, Mishra SK, Ohara H, Shimada K, et al. Dissemination of multidrug-resistant Klebsiella pneumoniae clinical isolates with various combinations of carbapenemases (NDM-1 and OXA-72) and 16S rRNA methylases (ArmA, RmtC and RmtF) in Nepal. International Journal of Antimicrobial Agents. 2013;42:372–4. doi:10.1016/j.ijantimicag.2013.06.014.

16. Leski TA, Taitt CR, Bangura U, Stockelman MG, Ansumana R, Cooper WH, et al. High prevalence of multidrug resistant Enterobacteriaceae isolated from outpatient urine samples but not the hospital environment in Bo, Sierra Leone. BMC Infectious Diseases. 2016;16:167. doi:10.1186/s12879-016-1495-1.

17. Leski T, Vora GJ, Taitt CR. Multidrug resistance determinants from NDM-1-producing Klebsiella pneumoniae in the USA. International Journal of Antimicrobial Agents. 2012;40:282–4. doi:10.1016/j.ijantimicag.2012.05.019.

18. Papp-Wallace KM, Endimiani A, Taracila MA, Bonomo RA. Carbapenems: past, present, and future. Antimicrobial Agents and Chemotherapy. 2011;55:4943–60. doi:10.1128/AAC.00296-11.

19. Kula B, Djordjevic G, Robinson JL. A systematic review: can one prescribe carbapenems to patients with IgE-mediated allergy to penicillins or cephalosporins? Clin Infect Dis. 2014;59:1113–22. doi:10.1093/cid/ciu587.

20. Wilson H, Török ME. Extended-spectrum β-lactamase-producing and carbapenemase-producing Enterobacteriaceae. Microbial Genomics 2018. doi:10.1099/mgen.0.000197.

21. Torok ME, Moran E, Cooke FJ. Oxford Handbook of Infectious Diseases and Microbiology. In: Oxford handbook of infectious diseases and microbiology. 2nd ed. [Oxford]: Oxford University Press; 2016.

22. P. Oduro. Prevalence of Carbapenemase Producing Enterobacteriaceae in Urinary Tract Infection Patients in Ghana. undefined. 2016.

23. Ssekatawa K, Byarugaba DK, Wampande E, Ejobi F. A systematic review: the current status of carbapenem resistance in East Africa. BMC Res Notes. 2018;11:629. doi:10.1186/s13104-018-3738-2.

24. Moyo SJ, Aboud S, Kasubi M, Lyamuya EF, Maselle SY. Antimicrobial resistance among producers and non-producers of extended spectrum beta-lactamases in urinary isolates at a tertiary Hospital in Tanzania. BMC Research Notes. 2010;3:348. doi:10.1186/1756-0500-3-348.

25. Warnes SL, Highmore CJ, Keevil CW. Horizontal transfer of antibiotic resistance genes on abiotic touch surfaces: implications for public health. Am Soc Microbiol. 2012;3:6–12. Am Soc Microbiol. 2012;3:6.

26. Moyo S, Haldorsen B, Aboud S, Blomberg B, Maselle SY, Sundsfjord A, et al. Identification of VIM-2-producing Pseudomonas aeruginosa from Tanzania is associated with sequence types 244 and 640 and the location of blaVIM-2 in a TniC integron. Antimicrobial Agents and Chemotherapy. 2015;59:682–5. doi:10.1128/AAC.01436-13.

27. Poirel L, Revathi G, Bernabeu S, Nordmann P. Detection of NDM-1-producing Klebsiella pneumoniae in Kenya. Antimicrobial Agents and Chemotherapy. 2011;55:934–6. doi:10.1128/AAC.01247-10.

28. Revathi G, Siu LK, Lu P-L, Huang L-Y. First report of NDM-1-producing Acinetobacter baumannii in East Africa. Int J Infect Dis. 2013;17:e1255–8. doi:10.1016/j.ijid.2013.07.016.

29. Perovic O, Ismail H, Quan V, Bamford C, Nana T, Chibabhai V, et al. Carbapenem-resistant Enterobacteriaceae in patients with bacteraemia at tertiary hospitals in South Africa, 2015 to 2018. European Journal of Clinical Microbiology & Infectious Diseases. 2020;39:1287–94. doi:10.1007/s10096-020-03845-4.

30. Adesanya OA, Igwe HA. Carbapenem-resistant Enterobacteriaceae (CRE) and gram-negative bacterial infections in south-west Nigeria: a retrospective epidemiological surveillance study. AIMS Public Health. 2020;7:804–15. doi:10.3934/publichealth.2020062.

31. Vasaikar SD, Hanise P, Abaver DT. Epidemiology, risk factors and molecular analysis of carbapenem-resistant Enterobacteriaceae (CRE) in Mthatha, Eastern Cape, South Africa. International Journal of Infectious Diseases. 2020;101:54. doi:10.1016/j.ijid.2020.09.173.

32. Thomas TSM, Duse AG. Epidemiology of carbapenem-resistant Enterobacteriaceae (CRE) and comparison of the phenotypic versus genotypic screening tests for the detection of carbapenemases at a tertiary level, academic hospital in Johannesburg, South Africa. Southern African Journal of Infectious Diseases. 2018:1–7. doi:10.1080/23120053.2018.1509184.

33. Adrian Brink, Jennifer Coetzee, Cornelis Clay, Craig Corcoran, Johan van Greune, J D Deetlefs, et al. The spread of carbapenem-resistant Enterobacteriaceae in South Africa: Risk factors for acquisition and prevention. South African Medical Journal. 2012;102:599–601.

34. Ngbede EO, Adekanmbi F, Poudel A, Kalalah A, Kelly P, Yang Y, et al. Concurrent Resistance to Carbapenem and Colistin Among Enterobacteriaceae Recovered From Human and Animal Sources in Nigeria Is Associated With Multiple Genetic Mechanisms. Front. Microbiol. 2021;12:740348. doi:10.3389/fmicb.2021.740348.

35. El Wartiti MA, Bahmani F-Z, Elouennass M, Benouda A. Prevalence of carbapenemase-producing enterobacteriaceae in a University Hospital in Rabat, Morocco: A 19-months prospective study. International Arabic Journal of Antimicrobial Agents. 2012;2:1–6. doi:10.3823/718.

36. Quansah E, Amoah Barnie P, Omane Acheampong D, Obiri-Yeboah D, Odarkor Mills R, Asmah E, et al. Geographical Distribution of β-Lactam Resistance among Klebsiella spp. from Selected Health Facilities in Ghana. Tropical Medicine and Infectious Disease 2019. doi:10.3390/tropicalmed4030117.

37. Codjoe FS, Donkor ES, Smith TJ, Miller K. Phenotypic and Genotypic Characterization of Carbapenem-Resistant Gram-Negative Bacilli Pathogens from Hospitals in Ghana. Microbial Drug Resistance. 2019;25:1449–57. doi:10.1089/mdr.2018.0278.

38. Cape Coast Teaching Hospital. Cape Coast Teaching Hospital:: 2018 Performance Report. Cape Coast; 2018.

39. Ghana Statistical Service. 2010 Population & Housing Census:: District analytical report. Cape Coast Municipality. Accra; 2010.

40. CLSI. Performance Standards for Antimicrobial Susceptibility Testing. 28th ed. Wayne, PA; 2018.

41. Huttner A, Harbarth S, Carlet J, Cosgrove S, Goossens H, Holmes A, et al. Antimicrobial resistance: a global view from the 2013 World Healthcare-Associated Infections Forum. Antimicrob Resist Infect Control. 2013;2:31. doi:10.1186/2047-2994-2-31.

42. Samuel S, Kayode O, Musa O, Nwigwe G, Aboderin A, Salami T, Taiwo S. Nosocomial infections and the challenges of control in developing countries. Afr J Clin Exp Microbiol. 2010;11:102–9.

43. Shaikh S, Fatima J, Shakil S, Rizvi SMD, Kamal MA. Antibiotic resistance and extended spectrum beta-lactamases: Types, epidemiology and treatment. Saudi Journal of Biological Sciences. 2015;22:90–101. doi:10.1016/j.sjbs.2014.08.002.

44. Jacoby GA, Munoz-Price LS. The new beta-lactamases. N Engl J Med. 2005;352:380–91. doi:10.1056/NEJMra041359.

45. Blomberg B, Jureen R, Manji KP, Tamim BS, Mwakagile DSM, Urassa WK, et al. High rate of fatal cases of pediatric septicemia caused by gram-negative bacteria with extended-spectrum beta-lactamases in Dar es Salaam, Tanzania. Journal of Clinical Microbiology. 2005;43:745–9. doi:10.1128/JCM.43.2.745-749.2005.

46. Codjoe FS. Detection and characterisation of carbapenem-resistant gram-negative bacilli infections in Ghana.

47. Feglo PK, Gbedema SY, Quay SNA, Adu-Sarkodie Y, Opoku-Okrah C. Occurrence, species distribution and antibiotic resistance of Proteus isolates: A case study at the Komfo Anokye Teaching Hospital (KATH) in Ghana. International Journal of Pharma Sciences and Research. 2010;1.

48. Sami H, Sultan A, Rizvi M, Khan F, Ahmad S, Shukla I, Khan H. Citrobacter as a uropathogen, its prevalence and antibiotics susceptibility pattern. CHRISMED J Health Res. 2017;4:23. doi:10.4103/2348-3334.196037.

49. Nordmann P, Cuzon G, Naas T. The real threat of Klebsiella pneumoniae carbapenemase-producing bacteria. The Lancet Infectious Diseases. 2009;9:228–36. doi:10.1016/S1473-3099(09)70054-4.

50. Donkor ES, Dayie NT, Badoe EV. Vaccination against pneumococcus in West Africa: perspectives and prospects. Int J Gen Med. 2013;6:757–64. doi:10.2147/IJGM.S45842.

51. Labi A-K, Obeng-Nkrumah N, Bjerrum S, Enweronu-Laryea C, Newman MJ. Neonatal bloodstream infections in a Ghanaian Tertiary Hospital: Are the current antibiotic recommendations adequate? BMC Infect Dis. 2016;16:598. doi:10.1186/s12879-016-1913-4.

52. Ogbolu DO, Daini OA, Ogunledun A, Alli AO, Webber MA. High levels of multidrug resistance in clinical isolates of Gram-negative pathogens from Nigeria. International Journal of Antimicrobial Agents. 2011;37:62–6. doi:10.1016/j.ijantimicag.2010.08.019.

53. Carroll M, Rangaiahagari A, Musabeyezu E, Singer D, Ogbuagu O. Five-Year Antimicrobial Susceptibility Trends Among Bacterial Isolates from a Tertiary Health-Care Facility in Kigali, Rwanda. The American Journal of Tropical Medicine and Hygiene. 2016;95:1277–83. doi:10.4269/ajtmh.16-0392.

54. Muluye D, Wondimeneh Y, Ferede G, Nega T, Adane K, Biadgo B, et al. Bacterial isolates and their antibiotic susceptibility patterns among patients with pus and/or wound discharge at Gondar university hospital. BMC Research Notes. 2014;7:619. doi:10.1186/1756-0500-7-619.

55. Mbanga J, Dube S, Munyanduki H. Prevalence and drug resistance in bacteria of the urinary tract infections in Bulawayo province, Zimbabwe. East Afr J Public Health. 2010;7:229–32.

56. Kumburu HH, Sonda T, Mmbaga BT, Alifrangis M, Lund O, Kibiki G, Aarestrup FM. Patterns of infections, aetiological agents and antimicrobial resistance at a tertiary care hospital in northern Tanzania. Tropical Medicine and International Health. 2017;22:454–64. doi:10.1111/tmi.12836.

57. Mshana SE, Matee M, Rweyemamu M. Antimicrobial resistance in human and animal pathogens in Zambia, Democratic Republic of Congo, Mozambique and Tanzania: an urgent need of a sustainable surveillance system. Annals of Clinical Microbiology and Antimicrobials. 2013;12:28. doi:10.1186/1476-0711-12-28.

58. Ocan M, Bwanga F, Bbosa GS, Bagenda D, Waako P, Ogwal-Okeng J, Obua C. Patterns and predictors of self-medication in northern Uganda. PLoS ONE. 2014;9:e92323. doi:10.1371/journal.pone.0092323.

59. Donkor ES, Tetteh-Quarcoo PB, Nartey P, Agyeman IO. Self-medication practices with antibiotics among tertiary level students in Accra, Ghana: a cross-sectional study. International Journal of Environmental Research and Public Health. 2012;9:3519–29. doi:10.3390/ijerph9103519.

60. Oduro-Mensah D, Obeng-Nkrumah N, Bonney EY, Oduro-Mensah E, Twum-Danso K, Osei YD, Sackey ST. Genetic characterization of TEM-type ESBL-associated antibacterial resistance in Enterobacteriaceae in a tertiary hospital in Ghana. Annals of Clinical Microbiology and Antimicrobials. 2016;15:29. doi:10.1186/s12941-016-0144-2.

61. Khanfar HS, Bindayna KM, Senok AC, Botta GA. Extended spectrum beta-lactamases (ESBL) in Escherichia coli and Klebsiella pneumoniae: trends in the hospital and community settings. J Infect Dev Ctries. 2009;3:295–9. doi:10.3855/jidc.127.

62. Ouedraogo A-S, Sanou M, Kissou A, Sanou S, Solaré H, Kaboré F, et al. High prevalence of extended-spectrum ß-lactamase producing enterobacteriaceae among clinical isolates in Burkina Faso. BMC Infectious Diseases. 2016;16:326. doi:10.1186/s12879-016-1655-3.

63. Bonnet R. Growing group of extended-spectrum beta-lactamases: the CTX-M enzymes. Antimicrobial Agents and Chemotherapy. 2004;48:1–14. doi:10.1128/AAC.48.1.1-14.2004.

64. Ehrhard I, Karaalp A-K, Hackel T, Höll G, Rodewald N, Reif U, et al. Prävalenzerhebung zum Vorkommen von Carbapenemase-Bildnern in sächsischen Kliniken. [Prevalence of carbapenemase-producing bacteria in hospitals in Saxony, Germany]. Bundesgesundheitsblatt - Gesundheitsforschung - Gesundheitsschutz. 2014;57:406–13. doi:10.1007/s00103-013-1914-z.

65. Qin S, Fu Y, Zhang Q, Qi H, Wen JG, Xu H, et al. High incidence and endemic spread of NDM-1-positive Enterobacteriaceae in Henan Province, China. Antimicrobial Agents and Chemotherapy. 2014;58:4275–82. doi:10.1128/AAC.02813-13.

66. Brink A, Coetzee J, Clay C, Corcoran C, van Greune J, Deetlefs JD, et al. The spread of carbapenem-resistant Enterobacteriaceae in South Africa: Risk factors for acquisition and prevention. South African Medical Journal. 2012;102:599–601.

